# Severe Acute Respiratory Syndrome Coronavirus-2 seropositivity in South-Central Uganda, during 2019 - 2021

**DOI:** 10.1101/2021.09.13.21263414

**Authors:** Charles Ssuuna, Ronald Moses Galiwango, Edward Nelson Kankaka, Joseph Kagaayi, Anthony Ndyanabo, Godfrey Kigozi, Gertrude Nakigozi, Tom Lutalo, Robert Ssekubugu, John Bosco Wasswa, Anthony Mayinja, Martina Cathy Nakibuuka, Samiri Jamiru, John Baptist Oketch, Edward Muwanga, Larry William Chang, Mary Kate Grabowski, Maria Wawer, Ronald Gray, Mark Anderson, Michael Stec, Gavin Cloherty, Oliver Laeyendecker, Steven James Reynolds, Thomas C. Quinn, David Serwadda

**Affiliations:** Rakai Health Sciences Program, Kalisizo, Uganda; Makerere University School of Public Health, Kampala, Uganda; Uganda Virus Research Institute, Entebbe, Uganda; Kyotera District Health Office, Kyotera District local government, Ministry of Health, Uganda; Department of International Health, Johns Hopkins Bloomberg School of Public Health, Baltimore, MD; Department of Epidemiology, Johns Hopkins Bloomberg School of Public Health, Baltimore, MD; Division of Infectious Disease, Division of Medicine, Johns Hopkins School of Medicine, Baltimore, MD; Abbott Laboratories, Abbott Diagnostics Division, Abbott Park, Illinois, USA; Division of Intramural Research, National Institute of Allergy and Infectious Diseases, National Institutes of Health, Bethesda, MD

**Keywords:** SARS-CoV-2 seroprevalence, healthcare workers, COVID-19, South-central Uganda

## Abstract

Globally, key subpopulations have a high risk of contracting SARS-CoV-2. In Uganda, limited access to personal protective equipment amidst lack of clarity on the extent of the community disease burden may exacerbate this situation.

We assessed SARS-CoV-2 antibody seroprevalence among high-risk sub-populations, including healthcare workers, persons within the general population previously reporting experiencing key COVID-19 like symptoms and archived plasma specimens collected prior to confirmation of COVID-19 in Uganda.

We collected venous blood from HCWs at selected health facilities and from population-cohort participants who reported specific COVID-19 like symptoms in a prior phone-based survey conducted during the first national lockdown (May-August 2020). Pre-lockdown plasma collected from individuals considered high risk for SARS-CoV-2 infection was retrieved. Specimens were tested for antibodies to SARS-CoV-2 using the CoronaChek™ rapid COVID-19 IgM/IgG lateral flow test assay. IgM only positive samples were confirmed using a chemiluminescent microparticle immunoassay (ARCHITECT AdviseDx SARS-CoV-2 IgM) which targets the spike. SARS-CoV-2 exposure was defined as either confirmed IgM, both IgM and IgG or sole IgG positivity.

The seroprevalence of antibodies to SARS-CoV-2 in HCWs was 21.1% [95%CI: 18.2-24.2]. Of the phone-based survey participants, 11.9% [95%CI: 8.0-16.8] had antibodies to SARS-CoV-2. Among 636 pre-lockdown plasma specimens, 1.7% [95%CI: 0.9-3.1] were reactive.

Findings suggest a high seroprevalence of antibodies to SARS-CoV-2 among HCWs and substantial exposure in persons presenting with specific COVID-19 like symptoms in the general population of South-central Uganda. Based on current limitations in serological test confirmation, it remains unclear whether pre-lockdown seropositivity implies prior SARS-CoV-2 exposure in Uganda.

## INTRODUCTION

It is over a year since SARS-CoV-2 emerged[1] as a global pandemic and as of the 2^nd^ of August 2021, nearly two hundred million cases were reported globally with >4,000,000 fatalities[2]. Transmission occurs by respiratory droplets, aerosols, and via fomites and is higher in confined or congested spaces[3]. SARS-CoV-2 infection can be asymptomatic[4] with estimates ranging from 5% – 80% while symptoms are largely nonspecific and include features of flu-like illness[5]. Diagnosis of asymptomatic and mild cases may be missed due to prioritization of screening/confirmatory tests for individuals with moderate to severe symptoms. However, asymptomatic and pre-symptomatic persons can be highly contagious and contribute greatly to epidemic spread[6, 7].

As of the 3^rd^ of August 2021, more than 94,000 cases with 2,710 deaths were documented in Uganda[2]. Community transmission is on the rise[8] despite earlier control measures that included a phased nationwide lockdown between March and August 2020[9]. The SARS-CoV-2 diagnostic testing landscape in Uganda prioritizes testing for symptomatic persons. It is unknown how many infected asymptomatic persons are missed due to this symptom-based testing approach and what impact this has on community transmission.

HCWs in particular are at a higher risk of contracting SARS-CoV-2[10, 11] and inadvertently transmitting it to their patients, some of whom may be immunocompromised. According to the World Health Organization (WHO), they account for 10% of the global SARS-CoV-2 burden[12]. This risk may be higher in countries like Uganda, due to shortage of Personal Protective Equipment (PPE) amidst unquantified community disease burden. Notably, several HCWs in Uganda have been infected and a number have died[13].

Due to the limited testing capacity, there are likely to be many undetected community infections fueling the epidemic. It is also unknown if SARS-CoV-2 importation or exposure in Uganda might have occurred earlier than the first (official) case reported on the 21^st^ of March 2020. We aimed at determining the prevalence of antibodies to SARS-CoV-2 among selected high-risk sub-populations in South-central Uganda, including HCWs, persons who previously reported specific COVID-19 like symptoms (fever, cough, loss of taste and smell) in the preceding 30 days, between May and August 2020. Additionally, we aimed at exploring the possibility of prior SARS-CoV-2 importation/exposure in South-Central Uganda before confirmation of the first (official) case on the 21^st^ of March 2020.

## METHODS

### Ethical approval

The study was approved by the Uganda Virus Research Institute’s Research Ethics Committee (Ref. GC/127/20/08/785), registered, and cleared by the Uganda National Council for Science and Technology (UNCST) (registration number HS878ES). Written informed consent was obtained from participants before blood specimens and other data were collected. Also, only archived pre-lockdown plasma specimens from Rakai Community Cohort Study (RCCS) participants that had provided prior consent for use of their blood specimens in future studies were retrieved to assess prior SARS-CoV-2 exposure in Uganda.

### Study design and setting

This study was cross-sectional and was conducted at the Rakai Health Sciences Program (RHSP) with participants recruited from within and outside the Rakai Community Cohort Study (RCCS) in four districts of South-central Uganda (Masaka, Kyotera, Rakai and Lyantonde). The RCCS is an open, population-based cohort in 40 communities in these districts with surveys conducted ∼ every 18 months among ∼ 23,000 adults, resident in fishing, agrarian, or peri-urban/trading community settings[14].

### Study population and sample size

A total of 980 participants including 753 HCWs and 227 individuals from the RCCS phone-based survey were recruited into the study. Participants from the cohort had previously reported experiencing COVID-19 like symptoms (fever, cough, loss of taste and/or loss of smell) in the preceding 30 days during an earlier phone-based survey conducted between May and August 2020. HCWs were identified from health facilities in the region, prioritizing high volume facilities located near the Uganda-Tanzania border or along the Kampala-Mutukula highway serving mobile persons who may be at higher risk of SARS-CoV-2 acquisition. At the selected health facilities, all available, willing HCWs were recruited into the study.

Additionally, we retrieved 636 archived plasma specimens collected between October 2019 and March 18^th^, 2020, before the first national lockdown took effect. Selected samples of persons living in RCCS communities close to the Tanzanian border and along the Kampala-Mutukula highway were considered to have a high risk for SARS-CoV-2 infection due to their high mobility and interaction with cross-border populations. They included traders/vendors, commercial sex work clients, fisher folks, bike (boda-boda) riders, truck drivers, mechanics, shopkeepers, and bar owners/workers.

### Sample / Data collection

Participants in the phone-based survey conducted during the first lockdown and reported having previously experienced at least one of the above COVID-like symptoms were contacted for participation in this study. Additionally, study field teams approached HCWs at selected health facilities for participation. Consenting participants provided 4mls of venous blood specimens while a short questionnaire was administered to HCWs to collect data on participant demographics, cadre, prior SARS-CoV-2 exposure, and PPE access/use. Plasma was frozen (−80**°**C) until laboratory analysis.

### Laboratory analysis

Frozen plasma was thawed and tested for antibodies to SARS-CoV-2 using the CoronaChek™ rapid COVID-19 IgM/IgG lateral flow test assay as per manufacturer’s instructions. This assay was previously validated with Ugandan samples, including 1077 pre-pandemic samples from the RCCS[15]. Low specificities of SARS-CoV-2 antibody assays have been reported, particularly from malaria endemic regions[16, 17]. Therefore, any sample that was solely IgM positive by CoronaChek™ was retested by the Abbott ARCHITECT AdviseDx SARS-CoV-2 IgM chemiluminescent microparticle immunoassay (CMIA) (Abbott, Chicago, IL).

### Data analysis

SARS-CoV-2 exposure was defined as either IgM confirmed by the ARCHITECT CMIA assay, both IgM and IgG or IgG sole positivity. Point prevalence and 95% confidence intervals were determined for each sub-group using the exact Clopper-Pearson method of calculating confidence intervals for binomial proportions.

## RESULTS

### Healthcare workers’ SARS-CoV-2 antibody test results

Most of the participants were female (64.54%) and were 25-34 years of age (31.6%). In the initial screening using the CoronaChek™, 30.8% (232/753) of HCWs had detectable SARS-CoV-2 antibodies irrespective of isotype class. Of these, 119 tested positive for IgM only, 102 for both IgM and IgG and 11 for IgG only. Of the initially 119 IgM only reactive samples, 46 were confirmed positive when re-tested using the ARCHITECT assay. The overall seroprevalence of SARS-CoV-2 antibodies among HCWs was 21.1% [95%CI: 18.2-24.2] (159/753). Majority (24/26) of the sampled health facilities had at least one healthcare worker who had antibodies to SARS-CoV-2. Seropositivity was highest among nurses and lowest among medical officers (**Table 1**).

**Table 1:** Factors associated with SARS-CoV-2 seropositivity among Healthcare workers.

| Sociodemographic characteristics |  | n (%) seropositive | <u>Univariate</u> |
| --- | --- | --- | --- |
|  |  | N=159 | Odds Ratio (95% CI) |
| Sex | Male | 61 (38.4) | 0.9 (0.6-1.2) |
|  | Female | 98 (61.6) | Ref |
| Age category | 15-24 | 33 (20.8) | 0.7 (0.4- 1.1) |
|  | 25-34 | 57 (35.8) | Ref. |
|  | 35-44 | 39 (24.5) | 0.8 (0.5-1.3) |
|  | 45-54 | 21 (13.2) | 1.1 (0.6-1.9) |
|  | 55+ | 9 (5.7) | 0.7 (0.3-1.5) |
| Cadre | Medical Officer | 3 (1.9) | 1.3 (0.4-5.1) |
|  | Clinical Officer | 8 (5.0) | 1.8 (0.7-4.3) |
|  | Nurse (all levels) | 57 (35.8) | Ref |
|  | Lab tech (all levels) | 16 (10.1) | 1.4 (0.8-2.7) |
|  | Other staff | 75 (47.2) | 1.0 (0.7-1.5) |

A total of 128 HCWs reported having undergone prior SARS-CoV-2 RT-PCR testing with 16 reporting a positive result. Of the 16 individuals, 8 had detectable antibodies to SARS-CoV-2. Out of the 128, a total of 108 HCWs reported previous negative RT-PCR results and 27% of these, subsequently tested antibody positive. Only face masks were reported to have been used by all HCWs who reported prior contact with a confirmed COVID-19 case. Despite reporting consistent use of face masks, 40% (63/156) of the HCWs reporting previous contact with a confirmed COVID-19 case had antibodies to SARS-CoV-2.

### Cohort participants’ SARS-CoV-2 antibody test results

Females comprised 69.1% and most participants were aged 35-44 years. Upon initial screening using the CoronaChek™, 16.3% of the participants (37/227) tested positive on IgM only, 2.2% (5/227) tested positive on IgG only whereas 6.6% (15/227) were positive on both IgM and IgG. Following retesting of the initially IgM only reactive samples using the ARCHITECT assay, 7/37 were confirmed positive. The overall seroprevalence of antibodies to SARS-CoV-2 in this population was 11.9% [95%CI: 8.0-16.8] (27/227). There was nearly no difference in seropositivity among HIV positive and negative participants (**Table 2**).

**Table 2:**
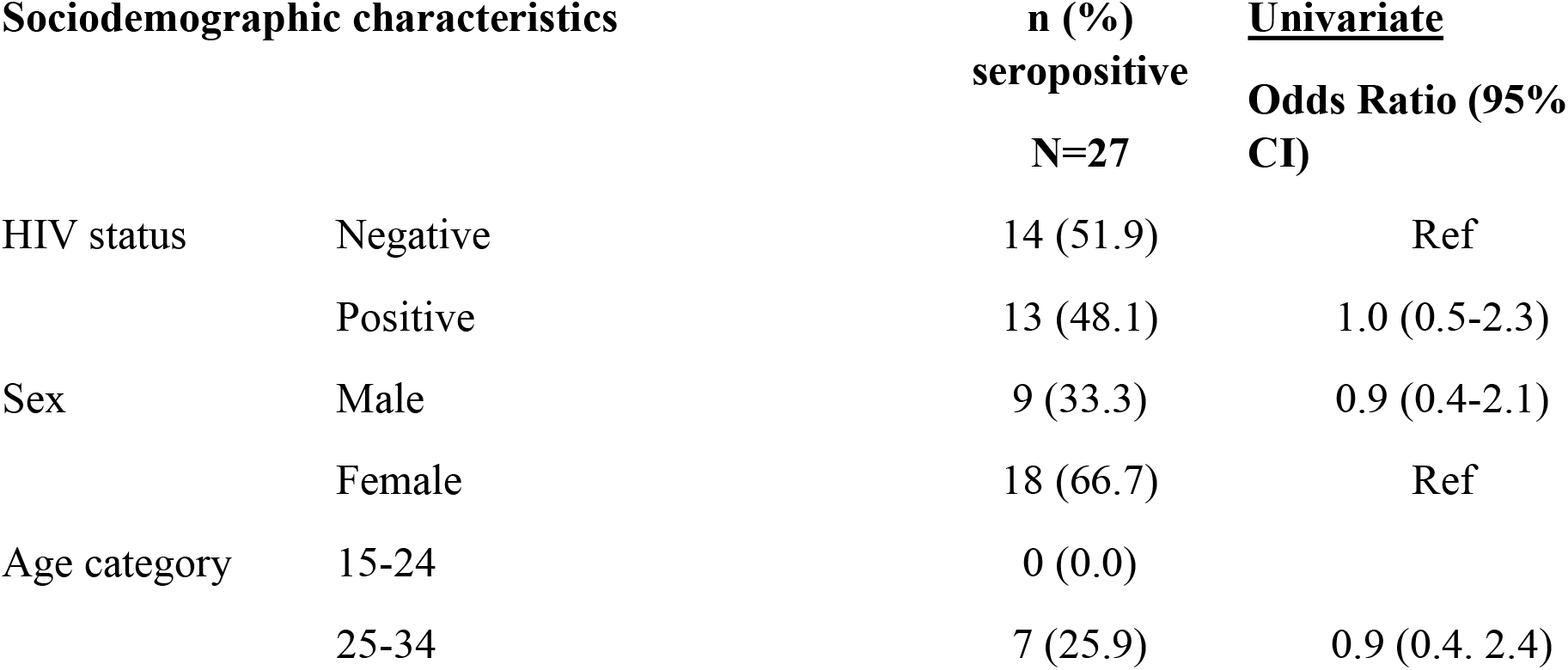
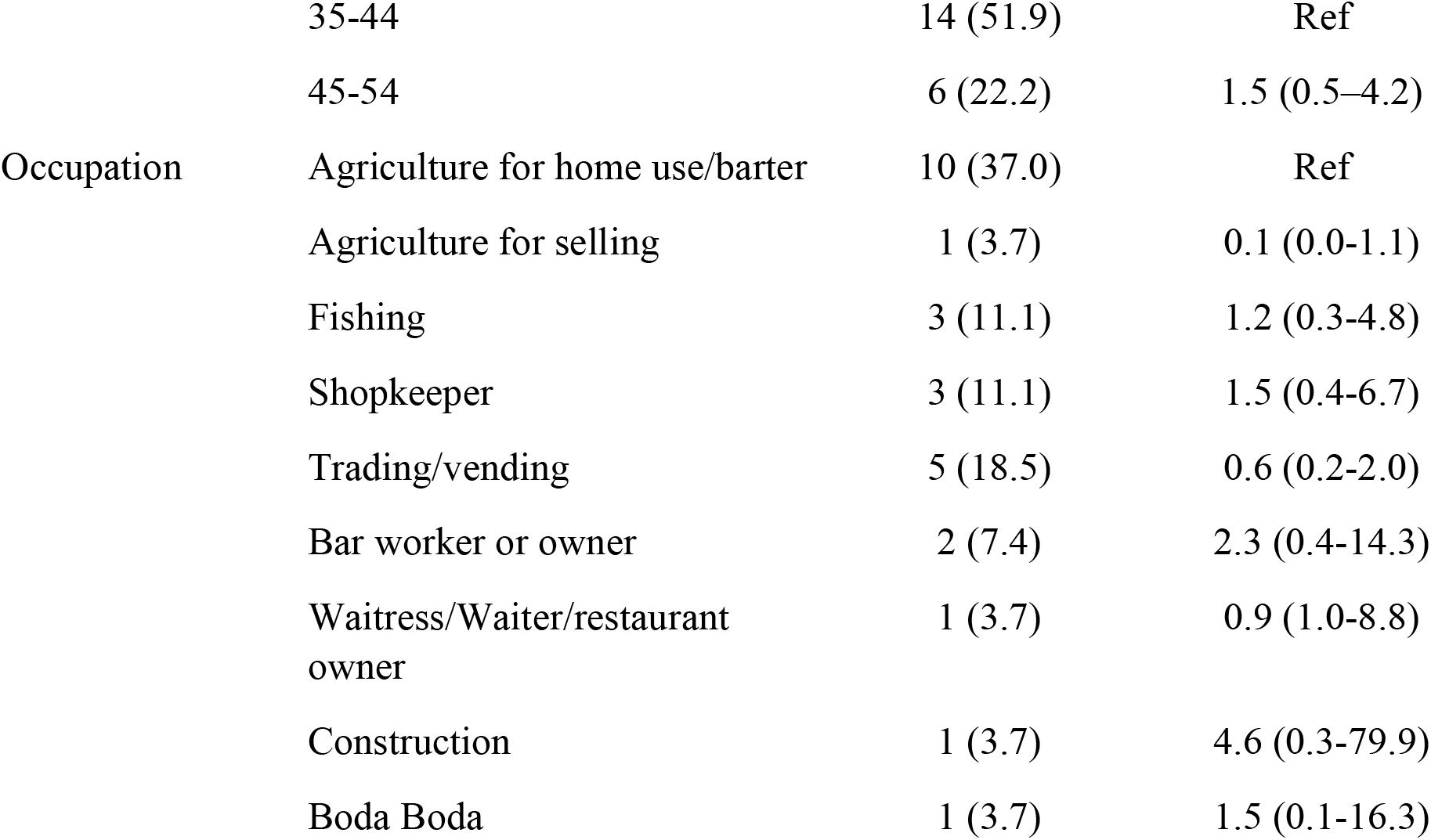
Factors associated with SARS-COV-2 seropositivity among phone-based survey participants.

### Pre-lockdown SARS-CoV-2 antibody test results

Upon initial screening using the CoronaChek™, 7% (47/363) of specimens had detectable antibodies to SARS-CoV-2 irrespective of isotype class. The majority (44) were IgM sole reactive, 2 were positive on IgG only whereas 1 reacted for both IgM and IgG. Out of the 44 IgM sole positive samples, 8 were confirmed following re-testing using the ARCHITECT assay. The overall seroprevalence of antibodies to SARS-CoV-2 in this sample category was 1.7% [95%CI: 0.9-3.1] (11/636).

## DISCUSSION

These findings suggest a relatively high SARS-CoV-2 seroprevalence among HCWs at almost all the selected health facilities (24/26) in South-central Uganda and substantial seroprevalence in persons previously reporting specific COVID-19 like symptoms within the general population. There was also potentially a spike in transmission a few weeks prior to this evaluation with predominance of IgM only antibodies in most of the participants.

There are challenges interpreting SARS-CoV-2 rapid serology in regions with high malaria endemicity as infection with *Plasmodium* species was shown to induce cross-reactive antibodies to carbohydrate epitopes on the SARS-CoV-2 spike protein[17, 18]. It is thus unclear whether seropositivity in pre-lockdown plasma specimens implies prior SARS-CoV-2 or other related coronavirus exposure or malaria in Uganda.

HCWs are minimally protected by face masks and only a few had accesses to other PPE (face shields, gowns, aprons etc.) and this, coupled with likelihood of improper face mask use or lack of N95-level protection, could explain the positive COVD-19 antibody results observed even among participants reporting face mask use. Several undetected cases among HCWs in this region is a potential driver of nosocomial spread. A moderate concordance between reported RT-PCR COVID-19 positives and antibody test outcome may reflect waning antibody levels as reported in several publications [19, 20].

## Data Availability

All data has been provided in the manuscript.

## Acknowledgements

**Source of funding:** This work was funded by the Government of Uganda through Makerere University Research and Innovations Fund (Grant number RIF/COVID/075, https://rif.mak.ac.ug/) and in part by the Division of Intramural Research, National Institute of Allergy, and Infectious Diseases (NIH, https://www.niaid.nih.gov/about/dir). The funders had no role in study design, data collection and interpretation, or the decision to submit the work for publication. **Personal assistance: Samples/Data collection**: RCCS field team, District Health Officers of Rakai, Kyotera, Lyantonde and Masaka districts, Prossy Namutebi, Wilson Bwanike; **Data management**: Damalie Nansimbi, Muhammed Mugerwa, Darix Ssebagala Kigozi

## Author contributions

**Protocol development:** Ronald M. Galiwango, David Serwadda, Edward N. Kankaka, Charles Ssuuna, Thomas C. Quinn, Kate M. Grabowski, Larry W. Chang, Steven J. Reynolds, Maria J. Wawer, Ronald H. Gray; **Study implementation:** Charles Ssuuna, Ronald M. Galiwango, David Serwadda, Robert Ssekubugu, John Bosco Wasswa, Anthony Ndyanabo, Edward Muwanga, Martina Cathy Nakibuuka, Samiri Jamiru, John Baptist Oketch, Anthony Mayinja Mark Anderson, Michael Stec, Gavin Cloherty; **Manuscript development:** Charles Ssuuna, David Serwadda, Ronald M. Galiwango, Edward Kankaka, Joseph Kagaayi, Godfrey Kigozi, Gertrude Nakigozi, Thomas C. Quinn, Kate M. Grabowski, Larry W. Chang, Steven J. Reynolds, Tom Lutalo, Maria J. Wawer, Ronald H. Gray, Oliver Laeyendecker

